# Tobacco, nicotine, and cannabis use and exposure in an Australian Indigenous population during pregnancy: A protocol to measure parental and foetal exposure and outcomes

**DOI:** 10.1101/2024.02.29.24303540

**Authors:** Angela Ratsch, Elizabeth A. Burmeister, Aunty Veronica Bird, Aunty Joyce Bonner, Uncle Glen Miller, Aunty Marj Speedy, Graham Douglas, Stevan Ober, Ann Woolcock, Sharly Blair (nee Murdoch), Min-Tz Weng, Jared A. Miles, Kathryn J. Steadman

## Abstract

**Background:** The Australian National Perinatal Data Collection collates all live and stillbirths from States and Territories in Australia. In that database, maternal cigarette smoking is noted twice (smoking <20 weeks gestation; smoking >20 weeks gestation). Cannabis use and other forms of nicotine use, for example vaping and nicotine replacement therapy, are nor reported. The 2021 report shows the rate of smoking for Australian Indigenous mothers was 42% compared with 11% for Australian non-Indigenous mothers. Evidence shows that Indigenous babies exposed to maternal smoking have a higher rate of adverse outcomes compared to non-Indigenous babies exposed to maternal smoking.

**Objectives:** The reasons for the differences in health outcome between Indigenous and non-Indigenous pregnancies exposed to tobacco and nicotine is unknown but will be explored in this project through a number of activities. Firstly, the patterns of parental and household tobacco, nicotine and cannabis use and exposure will be mapped during pregnancy. Secondly, a range of biological samples will be collected to enable the first determination of Australian Indigenous people’s nicotine and cannabis metabolism during pregnancy; this assessment will be informed by pharmacogenomic analysis. Thirdly, the pharmacokinetic and pharmacogenomic findings will be considered against maternal, placental, foetal and neonatal outcomes. Lastly, an assessment of population health literacy and risk perception related to tobacco, nicotine and cannabis products peri-pregnancy will be undertaken.

**Methods:** This is a community-driven, co-designed, prospective, mixed-method observational study with regional Queensland parents expecting an Australian Indigenous baby and their close house-hold contacts during the peri-gestational period. The research utilises a multi-pronged and multi-disciplinary approach to explore interlinked objectives.

**Results:** A sample of 80 mothers expecting an Australian Indigenous baby will be recruited. This sample size will allow estimation of at least 90% sensitivity and specificity for the screening tool which maps the patterns of tobacco and nicotine use and exposure versus urinary cotinine with 95% CI within ±7% of the point estimate. The sample size required for other aspects of the research is less (pharmacokinetic and genomic n=50, and the placental aspects n=40), however from all 80 mothers, all samples will be collected.

**Conclusions:** Results will be reported using the STROBE guidelines for observational studies.

**Forward:** We acknowledge the Traditional Custodians, the Butchulla people, of the lands and waters upon which this research is conducted. We acknowledge their continuing connections to country and pay our respects to Elders past, present and emerging.

Notation: In this document, the terms Aboriginal and Torres Strait Islander and Indigenous are used interchangeably for Australia’s First Nations People. No disrespect is intended, and we acknowledge the rich cultural diversity of the groups of peoples that are the Traditional Custodians of the land with which they identify and with whom they share a connection and ancestry.

## Introduction

In 1957, Simpson [1] reported a dose-response association between maternal smoking and premature birth. This observation resulted in a world-wide research agenda focusing on the impact of maternal smoking in pregnancy, the findings of which indicate that maternal tobacco smoking, and exposure to the products of tobacco combustion (i.e., secondhand smoke exposure) are the leading modifiable risk behaviours associated with adverse maternal and neonatal outcomes. Maternal exposure increases the risk for miscarriage, ectopic pregnancy, antepartum bleeding, placental abruption, placenta previa, postpartum haemorrhage and alters maternal thyroid function [2, 3]. Counterintuitively, smoking in pregnancy is associated with decreased hypertensive disorders of pregnancy [4, 5]. For the foetus, maternal exposure increases the risk of stillbirth, premature birth and lower birthweight [6] as well as increases the risk for a number of congenital abnormalities [7, 8]. Longer-term, offspring exposed in-utero to maternal smoking have decreased cognitive achievement [9] and an increased risk for the development of attention-deficit/hyperactivity disorder (ADHD) [10].

In Australia, these adverse pregnancy and foetal findings have ensured a prenatal focus on maternal smoking behaviour with the mother’s cigarette smoking status obtained during antenatal assessment and recorded in the National Perinatal Data Collection [11] twice across the nine months of pregnancy (once < 20 weeks gestation, and once >20 weeks gestation). In that database, the 2021 rate of smoking by Australian non-Indigenous expectant mothers was reported as 11% compared with 42% by Australian Indigenous expectant mothers [12]. However, the maternal use of other tobacco and nicotine products including e-cigarettes, hookahs, chop-chop tobacco, nicotine tooth cleaning powder, chewing tobacco, and nicotine spray, mist, lozenges, gum and patches, and cannabis is not collected. In addition, this focus on maternal cigarette smoking fails to recognise second- and third-hand maternal nicotine vape and tobacco and cannabis smoke exposure, i.e., the impact from paternal and household tobacco, nicotine and cannabis exposure on maternal and foetal outcomes is overlooked.

This assessment gap results in maternal and foetal tobacco, nicotine and cannabis exposure misclassification and ramifications in the planning of care, and in the estimation of adverse maternal, placental, foetal and neonatal outcomes from tobacco, nicotine and cannabis exposure. Nevertheless, the literature indicates that Indigenous babies exposed to maternal smoking have a higher rate of adverse outcomes compared to non-Indigenous babies exposed to maternal smoking, for example, after adjusting for maternal age and other factors, smoking in pregnancy is attributable to 22% of pre-term Indigenous births compared with 5% for non-Indigenous births [13]. Gestational age impacts birthweight, and for Indigenous mothers who smoked in the first 20 weeks of pregnancy, the risk of a lower birthweight baby was 1.8 (or about 80% higher risk) than among Indigenous mothers who did not smoke in the first 20 weeks. For non-Indigenous mothers who smoked in pregnancy, this risk was 1.3 (or 30% higher risk) [14].

The other significant foetal outcome attributable to maternal smoking is stillbirth. In Australia, stillbirth is defined as foetal death prior to birth of the baby at 20 weeks gestation or more, and/or weighing 400 grams or more [15]. In 2020, the overall Australian stillbirth rate was 7.7/1000 births, with the rate for Indigenous mothers being 11.9/1000 compared with 7.4/1000 for non-Indigenous mothers. Smoking in pregnancy is a risk factor for stillbirth; for women who smoked in pregnancy, the stillbirth rate was 12.8 stillbirths/1000 births compared to 6.9 stillbirths/1000 births for mothers who did not smoke [15]. Sub-category analysis of smoking and stillbirth for Indigenous mothers compared to non-Indigenous mothers is not published.

The reasons for the differences in pregnancy outcome between Indigenous and non-Indigenous pregnancies exposed to tobacco and nicotine are unknown but will be explored in this project through a number of activities.

## Background

### Nicotine: pregnancy health and the developing human

Currently, tobacco assessment in pregnancy is focused on ‘smoking’. This emphasis overlooks the absorption of the pharmacologically active, dose-dependent, potentially lethal component of tobacco-which is nicotine-from other sources [16]. This study is premised on some of the actions of nicotine. In brief, nicotine binds with and activates nicotinic acetylcholine receptors (nAChR) in central and peripheral neuronal and non-neuronal tissue, at neuromuscular junctions, and in the adrenal medulla [17]. Receptor type and individual variability, including genetics and pregnancy, result in receptor up-regulation or desensitization [16, 18] and the release of neurotransmitters including acetylcholine, norepinephrine, dopamine, serotonin, vasopressin, beta-endorphin and adreno-corticotropic-hormone, thus impacting the vasculature and producing vasoconstriction, increasing heart rate and blood pressure [19].

In pregnancy, nicotine acts both directly on nAChRs in the developing placenta, reducing the number of nAChR receptors [20], and alters placental morphology and vascularity [21]. Nicotine also readily crosses the placenta and stimulates nAChRs in the developing foetus impacting foetal systems, albeit in an immature physiology [22]. During early foetal development (4-6 weeks of gestation), nAChRs emerge and begin to form connections throughout the body including the brain [23], with construction of axons and synaptic connections in the brain continuing after birth into childhood, adolescence and young adulthood [24]. In the foetal brain, nicotine exposure results in accelerated cell development relative to tissue and organ age, that is, there are fewer cells correctly developed for their stage and age [25]. Following nicotine exposure, changes in nAChRs and neural plasticity result in both a deficit in the number of neurons in the foetal brain and synaptic level damage to the respiratory center, with equivalent damage to the adrenal glands [26]. Foetal nicotine exposure ultimately results in a dampened response to hypoxic episodes [27]. These physiological changes are important in understanding links between maternal nicotine exposure and foetal outcomes, for example, stillbirth and Sudden Infant Death Syndrome (SIDS).

### Nicotine metabolism and excretion

Nicotine has a half-life of about two hours and is metabolised via the CYP2A6 pathway primarily in the liver, with the brain, kidneys and lungs providing minor sites [28]. This short half-life produces large nicotine serum plasma fluctuations and poses challenges for intra- and inter-person comparisons of exposure, consumption and effect. However, cotinine (the main tobacco and nicotine metabolite) has a half-life of approximately 17 hours and serum concentrations 10-fold higher than nicotine, providing a more stable biomarker of tobacco and nicotine exposure [29]. Cotinine begins to metabolise after approximately 16 hours into trans-3’-hydroxycotinine (3-OH-cotinine), nornicotine, nicotine glucuronide and nicotine-N-oxide [18]. A more accurate assessment of tobacco and nicotine exposure is achieved by measuring serum tobacco and nicotine and metabolite concentrations (total nicotine equivalents – TNE), as opposed to measuring only nicotine concentration [30].

During pregnancy, nicotine metabolism is impacted both by individual variability [16] and the changes created by pregnancy. In the expectant mother, there is a significant induction of CYP2A6 activity which increases plasma clearances of nicotine by 60% and cotinine by 140%, in addition, there is an almost 50% reduction in cotinine half-life (down from 17 hours to 9 hours [31]). This is important in the consideration of tobacco and nicotine use in pregnancy as decreases in nicotine and cotinine measurements in late pregnancy compared with pre-pregnancy or early pregnancy may not necessarily indicate a decrease in tobacco and nicotine exposure, but rather the more rapid metabolism of nicotine [32]. In the foetus and neonate, the immature and undeveloped CYP2A6 pathway decreases their ability to metabolise nicotine and results in a much longer plasma nicotine half-life than adults (11.2 hours compared to 2 hours) whereas cotinine elimination is similar to that of adults (16.3 hour half-life compared with 17 hours) [33].

CYP2A6, the major nicotine-oxidising enzyme, is measurable using the nicotine metabolite ratio (NMR; 3`′`hydroxycotinine:cotinine) and is a biomarker of nicotine clearance [16]. However, NMR is highly heritable (∼80%) varying with ethnicity, in part due to CYP2A6 variants. Variation in CYP2A6 genes has not been characterised in Australian Indigenous populations and may contribute to increased/decreased pregnancy risk from tobacco and nicotine exposure.

Tobacco, nicotine and the metabolites are rapidly excreted by the kidneys with the rate dependent upon urinary pH, where increased urine alkalinity decreases excretion [34]. Urinary excretion is further impacted in pregnancy due to elevated creatinine potentially leading to fluctuations in nicotine and its metabolites [35]. Thus, serum provides a measure of exposure and the recency of that exposure, while urine analysis provides a measure of tobacco and nicotine metabolism and excretion.

## Materials and Methods

### Aim and Objectives

The aim of this study is to develop a foundation from which approaches to tobacco and nicotine assessment, health literacy, tobacco and nicotine cessation, and maternal and neonatal health care delivery for families expecting an Australian Indigenous baby is informed by contemporary evidence. Community input on the design suggested that cannabis use (with and without tobacco) is also prevalent and continues throughout pregnancy, thus cannabis use and exposure have been included.

The objectives are to:

1. Accurately describe parental and close household contact(s) peri-gestational patterns of use and exposure of tobacco, nicotine and cannabis products through the creation and use of a validated assessment tool.
2. Determine the pharmacokinetic and pharmacogenomic impacts and outcomes of tobacco, nicotine and cannabis exposure. This study will be the first to establish the metabolism of tobacco, nicotine and cannabis during pregnancy by using NMR as a biomarker of individual differences in nicotine and cannabis metabolism in a parental Australian Indigenous population.
3. Describe maternal, paternal, placental, foetal and neonatal outcomes according to the use and exposure to tobacco, nicotine and cannabis products and biochemical and genomic analysis.
4. Describe the influences and barriers to cessation for pregnant Australian Indigenous families or close household contacts to reduce or cease tobacco, nicotine and/or cannabis use in pregnancy.

### Research Governance

This project is centred around the local Australian Indigenous population in the Fraser Coast area (Queensland, Australia). The research will be conducted primarily from Galangoor Duwalami Primary Healthcare Service (the local Aboriginal and Torres Strait Islander Primary Health Service in Hervey Bay and Maryborough) and at the Hervey Bay and Maryborough Hospitals within the Wide Bay Hospital and Health Service (WBHHS).

Galangoor Duwalami Primary Healthcare Service were consulted in regard to which Community members would best represent the Traditional Owners of the land where this research is to be conducted. A combined leadership group was formed which included the Butchulla Aboriginal Corporation and Butchulla Men’s Business Association and this group advised, directed and oversaw the conversations around this research from its inception and arranged for discussions with the appropriate Community members. Under the Butchulla Aboriginal Corporation’s Rule Book, the principal Objective of the corporation is to: ‘Relieve poverty and disadvantage of the Butchulla People through the advancement of education, health, social or public welfare, and culture’. Accordingly, the Elders have approved this proposal and provided a Butchulla name for the project – *Ngabang* (mother), *Walbai* (baby), *Babun* (father). The project logo (Figure 1) reflects the aspirations of the Butchulla people for this project, which are to strive for a *healthy pregnancy*, which results in a *healthy family*, which maintains a *healthy culture*.

**Figure 1.**
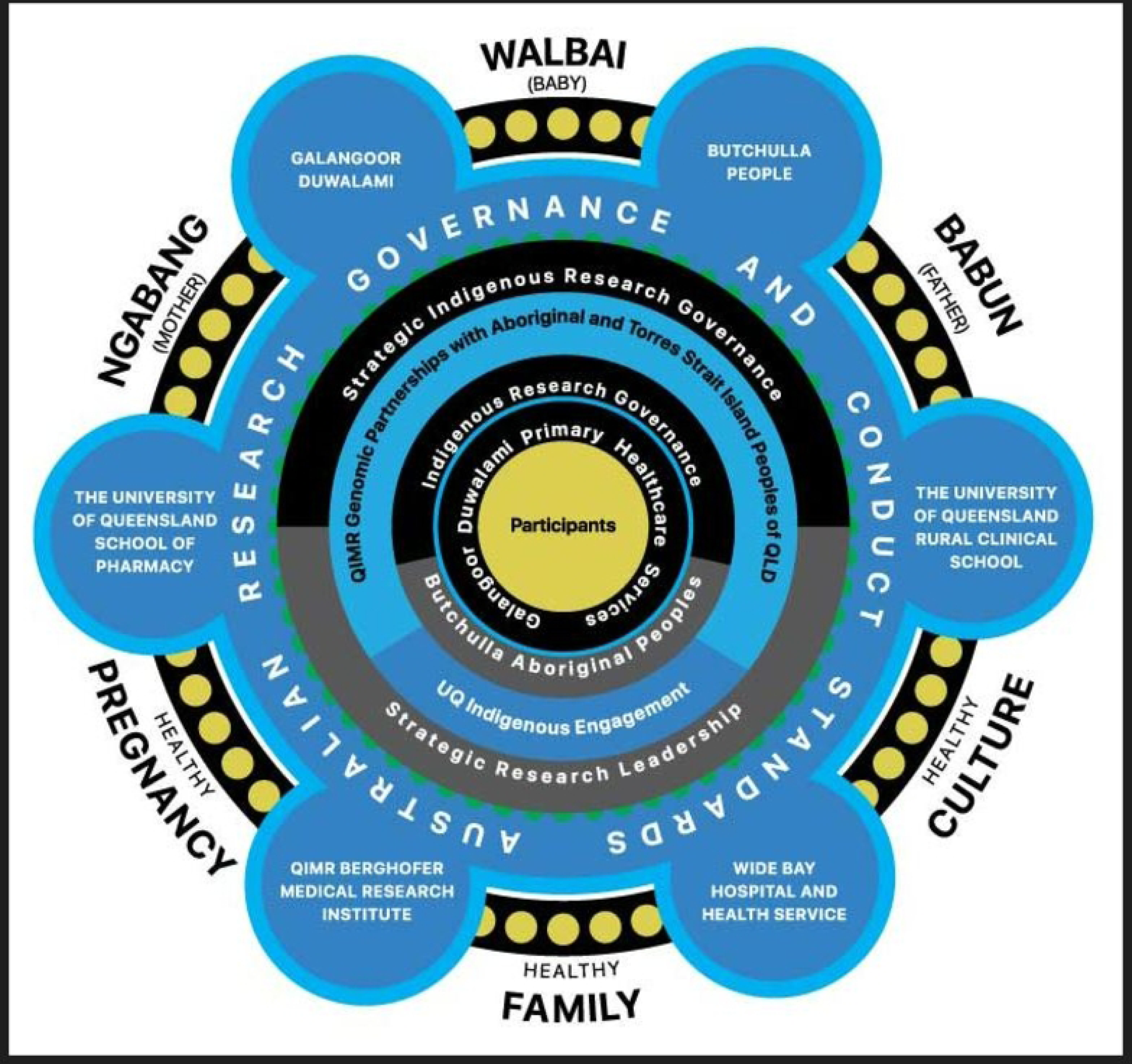
Project logo.

The logo also recognises the governance of the project by the Butchulla people, Galangoor Duwalami Primary Healthcare Service, the various Australian research guidelines, and the collaborative nature of the project with other health providers and research-intensive organisations and academic partners. The project documentation, and staff and participant shirts and onesies carry this logo (Figure 2 and 3)

**Figure 2.**
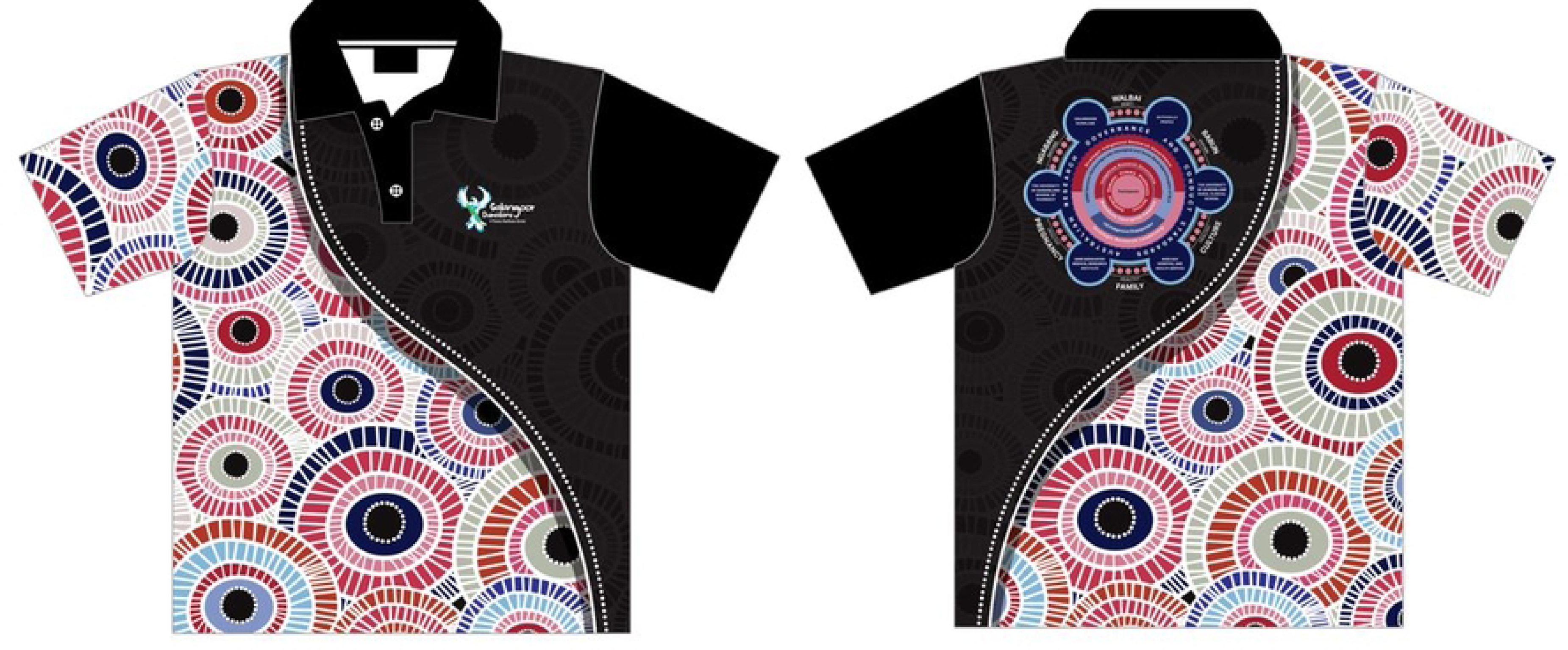
Participant shirt.

**Figure 3.**
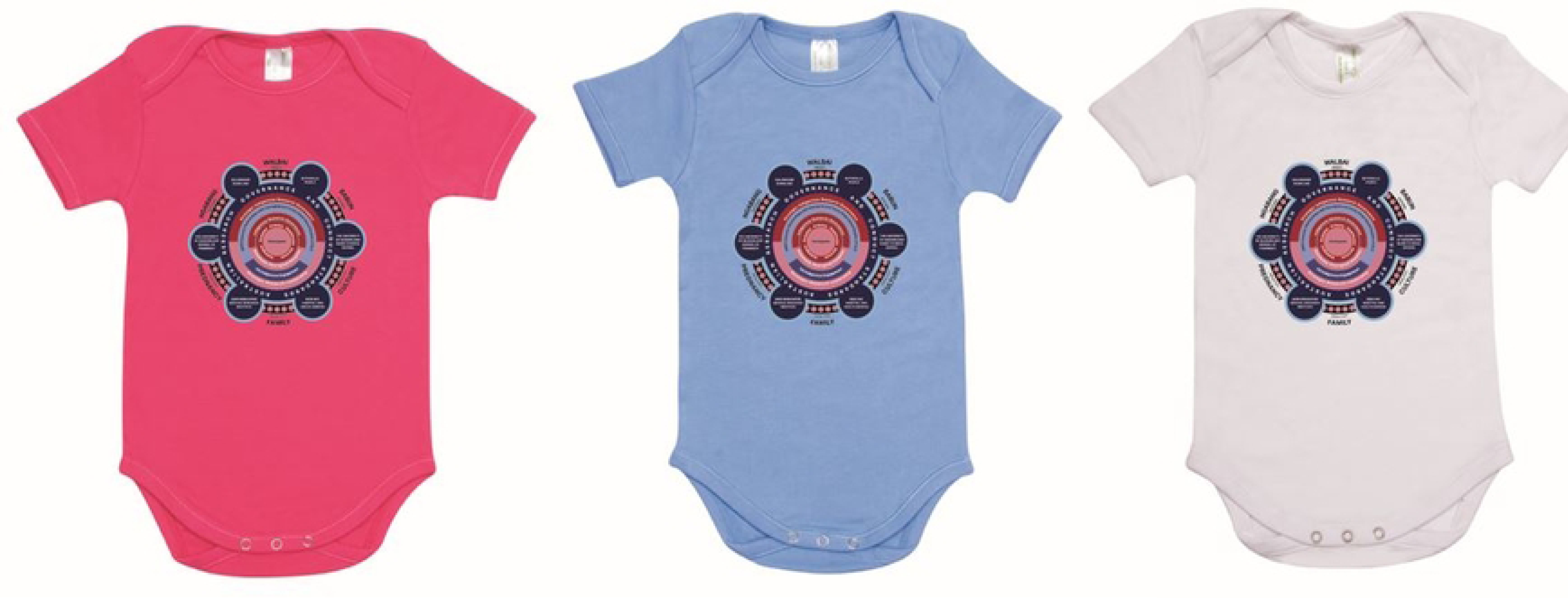
Participant onesie.

### Research context

Galangoor Duwalami Primary Healthcare Service generally maintain the sole care of the pregnant women until approximately 20 weeks gestation (unless they have a high-risk pregnancy). The mother then moves to a shared care model with the Hervey Bay and Maryborough Hospitals. In this project, data is collected across the entire pregnancy. The preferred site of data collection from Galangoor Duwalami Primary Healthcare Service patients will be at Galangoor Duwalami Primary Healthcare Service for as long as possible during the pregnancy, however some data collection will need to occur at antenatal visits at the Hervey Bay or Maryborough Hospitals. At birth (mostly likely at the Hervey Bay Hospital), biological samples will be collected, and birthing data will be collected on the participant’s discharge from hospital.

### Inclusion and exclusion criteria and consent

Potential participants who meet the following criteria will be invited to enrol in the study: pregnant mothers, aged 15 years or older at the time of enrollment, able to understand English, able to provide informed consent, and who self-identify as being pregnant with an Australian Indigenous baby. In accordance with the value of Respect outlined in the National Statement of Ethical Conduct in Human Research [36], all potential maternal participants meeting the inclusion criteria will be carefully considered by the clinic midwife or healthcare medical officer to ensure the healthcare staff believe that potential enrolment is in the best interest of the participant at this point in their pregnancy. A list of suitable potential maternal participants will then be provided to the researchers; only those participants will be provided with research information.

The project will strive to enrol the family unit i.e., the mother, and the foetus, and the biological father, or non-biological parent partner (the parents), or one close household contact. For example, if an expectant mother lives with her sister or aunty or grandmother but there is no biological father in the family at that time, or there is a partner (of either gender) and that close family contact attends the antenatal visits with the maternal participant, that person will be invited to participate with the expectant mother. Family members do not have to participate; however, ‘family’ is central to Australian Indigenous people, and enrolling participants who constitute the cultural norm of family will be especially important in the translation of findings to the participant and the community.

Consent: Participants can choose to enrol in any or all aspects of the data and sample collection, and they can withdraw from any or all aspects of the research at any time. The consent process enables participants to choose to have their individual results provided back to them at the completion of the project; for any or all samples to returned to the participant on completion of this project; or for the de-identified samples and de-identified data to be retained for ethically approved un-identified projects in the future. All enrolled participants will receive a participant shirt (Figure 2), and liveborn newborns will receive a project onesie (Figure 3).

A subset of information-rich tobacco, nicotine and/or cannabis use/exposed participants will be enrolled into the separate male and female qualitative aspects of the study until saturation is reached (∼30 participants). Data will be collected in response to a range of trigger questions and survey questions. Participants enrolled in the qualitative data collection will receive a $50 grocery voucher in acknowledgement of their time to the project.

### Tobacco, nicotine and cannabis use and exposure data and biological sample collection

The quantitative data and sample collection for Objectives 1-3 is designed to measure maternal tobacco, nicotine and cannabis use, and maternal and foetal exposure, metabolism and excretion at varying times throughout the pregnancy and assess the findings against the maternal and foetal outcomes. At each antenatal appointment and at birth, tobacco, nicotine and cannabis use and exposure information will collected on a ***N****ic**OTI**ne, tobacco, and **C**annabis use and **E**xposure (**NOTICE**) Assessment Tool* (Figure 4 and see Box 1 NOTICE Notes).

**Figure 4.**
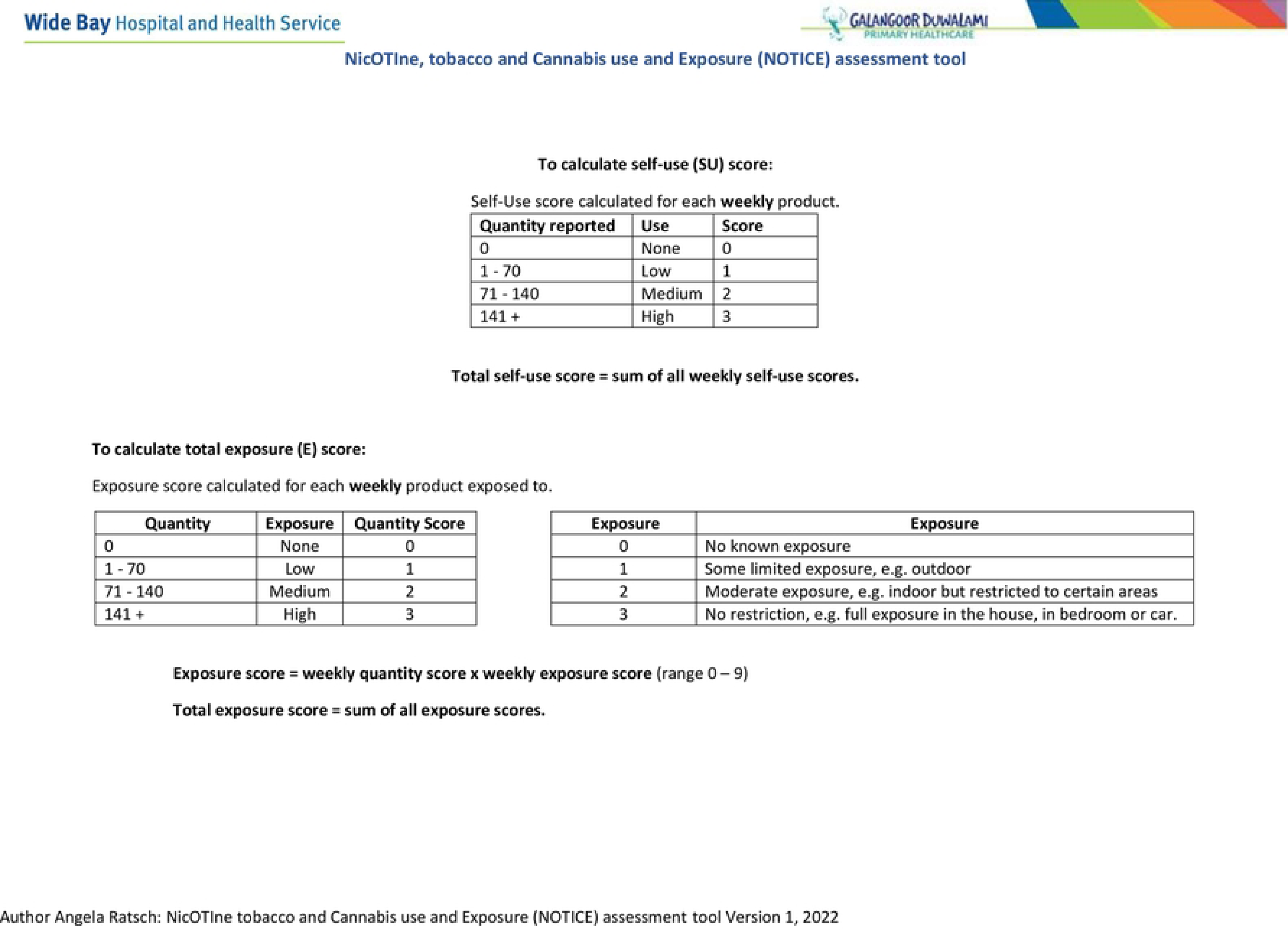
*NOTICE:* NicOTIne, tobacco, and Cannabis use and Exposure (NOTICE) Assessment Tool.

#### Box 1

**NicOTIne tobacco, and Cannabis use and Exposure (NOTICE)**

##### Assessment Tool – Notes

The **N**ic**OTI**ne tobacco, and **C**annabis use and **E**xposure (**NOTICE**) assessment tool (Figure 4) has been developed by Angela Ratsch for this study to enable a comprehensive assessment of the use of smoked and smokeless tobacco and cannabis products as well as nicotine containing products. The tool also allows for the detailing of second-hand exposure, and is used in this protocol to inform the use and exposure of all products throughout the pregnancy. The recorded information facilitates a scoring method to determine the level of self-reported use and exposure to tobacco, nicotine and cannabis products (scored for the day of assessment, previous 24 hours, and the previous week).

A self-use weekly score will be calculated for each assessment completed. Scoring for self-use will be categorized for each product as: 0 - no use; 1 – low use (1 to 70 combusted or inhaled products); 2 – medium use (71-140 combusted or inhaled products); 3 – high use (141 + combusted or inhaled products). A self-use score will be calculated for each weekly product combusted or inhaled with the sum of these scores producing a total weekly self-use score with a range of 0-30. An exposure score will be calculated for each weekly product exposed to by multiplying the quantity exposed to score by the exposure. Exposure is categorized as 0 - no exposure; 1-some limited exposure; 2 -moderate exposure or 3 - unrestricted exposure. A total weekly exposure score is then calculated by summing all the exposure scores with a range of 0-99 for each assessment completed.

The assessment takes about 3 minutes to complete and will be conducted at each antenatal visit and immediately prior to birth.

##### Exhaled breathe carbon monoxide (CO) assessment

CO as a point of care (POC) assessment provides indicative information about combusted tobacco/cannabis use up to 24 hours previously. Exhaled CO monitors (Smokerlyser®) provide a CO ppm reading ranging between 0 and 30 and a percentage of oxygen replaced with CO on haemoglobin (%COHb) with higher readings indicating higher levels of combusted tobacco/cannabis use. The same device can measure foetal exposure as foetal carboxyhaemoglobin (%FCOHb). Readings for the level of CO ppm are categorized as:

- green—for no and general environmental combusted tobacco/cannabis exposure, indicated by CO <3 ppm for pregnant women, and CO <6 ppm for non-pregnant adults;
- orange—moderate combusted tobacco/cannabis exposure, indicated by CO 4-6 ppm for pregnant women, and CO 7-10 ppm in non-pregnant adults and;
- red—heavy combusted tobacco/cannabis exposure, indicated by CO >6 ppm for pregnant women, and CO >10 ppm in non-pregnant adults.

The assessment takes about 1 minute to complete and will be conducted at each antenatal visit and immediately prior to birth.

##### Saliva cotinine assessment

The use of non-combusted tobacco products and nicotine substances, for example, chewed tobacco, e-cigarettes, nicotine patches and gum, does not create CO. Instead, cotinine saliva assessment (Oz Drug Tests®), as a POC measurement, can detect cotinine (the major metabolite of nicotine metabolism). The test has a cotinine cut-off value of 10 ng/mL and the expected window of detection for cotinine in saliva at 10 ng/mL is expected to be up to 2-3 days after the last nicotine use. The assessment takes about 3 minutes to complete and will be conducted at each antenatal visit and immediately prior to birth.

Procedure: The CO assessment will be conducted first, and if the CO level is within the yellow or red zone (high levels), a saliva test will not be conducted as these CO levels are indicative of combusted tobacco use and negates the requirement for saliva testing.

### Sample collection process

#### Mother

At the first antenatal appointment, maternal urine, CO and saliva samples will be collected on a NOTICE Tool.

If the CO is within the orange or red zones, a cotinine saliva test will not be conducted. Also, during this first appointment, or when maternal bloods are being collected as part of standard care, maternal vein blood will be collected for this project.

- At each follow-up antenatal visit, maternal urine, CO and saliva (if indicated) will be collected and a NOTICE assessment completed.

#### Biological father

- At the first antenatal appointment, if the biological father is Indigenous, paternal vein blood, urine and semen will be collected and CO, saliva (if indicated), and a NOTICE assessment completed.
- At each follow-up visit, the biologically paternal CO and saliva (if indicated) samples will be measured and a NOTICE assessment completed.

#### Non-biological parent, partner or close household member

- At the first antenatal visit, the non-biological parent partner or close household contact will have a CO and saliva (if indicated) measured and a NOTICE assessment completed and these same tests will be conducted at each visit.

#### Unaccompanied mother

- If the biological father, partner or household member is not present, the expectant mother will report her exposure to the partner’s and/or household member’s combusted tobacco or heated nicotine (e-cigarette) as second-hand exposure on the NOTICE assessment tool.

#### At birth

- On presentation for birth, mothers will have a venous blood sample, urine, CO and saliva samples (if indicated) collected and a NOTICE assessment will be completed.
- At spontaneous or artificial rupture of membranes or caesarean birth, amniotic fluid will be collected.
- Following birth and the separation of the placenta from the mother and neonate, arterial and venous cord bloods will be collected from the placenta.
- The placenta will be weighed in grams, and measured in centimetres at two points, the widest and narrowest to estimate the area of the placenta. Placental photographs of the maternal and foetal side and the cord will be obtained.
- Placental samples for macro, micro-morphological and genomic examination will be obtained, rinsed in a solution of phosphate buffered saline and distilled water and then placed in a solution of RNA later. The placenta sent for standard histology.

#### Postnatal

- Following birth, neonates will have a Day 0-1 urine and meconium collected. If the neonate remains in hospital after Day 1, a further urine sample will be collected. The urine will be collected with the use of a standard adhesive urine collection devise, and meconium will be collected from the nappy.
- Colostrum/breast milk will be collected when available. As active participants in the knowledge generation from this research, mothers will be encouraged to obtain their own colostrum/breast milk sample by hand/pump expression.

### Clinical information

Data contained in the Galangoor Duwalami Primary Healthcare Service and Hospital record including demographic, maternal, and neonatal data will be included in the research database. Pregnancy, labour and birthing information will be collected from the Queensland Health Perinatal Record following birth.

### Biological sample analysis

After labelling, participant samples will be stored in the appropriate solution and/or temperature control manner. Placental samples in RNA later will be transferred to the University of the Sunshine Coast for storage at −80°C. Other samples (excluding the whole placenta in formalin) will be transferred to the local Sullivan and Nicolaides Pathology at standard intervals. Sullivan and Nicolaides will then transfer directly to The University of Queensland for storage at −80°C. The placenta in formalin will be transferred to Queensland Pathology. The scientists undertaking the biological examinations and analysis will be blinded to the self-reported tobacco, nicotine and cannabis status of the participant.

Pregnancy elevates creatinine levels, potentially leading to fluctuations in the levels of nicotine and its metabolites and influencing cannabinoid levels [35]. To ensure the standardisation of these levels in urine samples during pregnancy, a creatinine analysis will be conducted. The biological samples will be initially analysed for tobacco, nicotine and their metabolites and cannabinoid concentration in ng/mg creatinine or nmol/mg creatinine for urine samples and ng/mL for other biological samples using standard procedures. Following this, the samples will undergo tobacco, nicotine and cannabinoid genomic assessment in relation to tobacco, nicotine and cannabis pharmacokinetics, and other examinations including thyroid antibodies, transthyretin and sENG levels and a genomic-wide analysis of tobacco nicotine and cannabis induced alterations will be conducted. In addition, the colostrum/breast milk microbiome will be considered for the impact of tobacco, nicotine and cannabis exposure.

### Qualitative data collection

Information-rich participants will be invited to take part in a face-to-face interview and survey. The interview and survey will be in the form of a yarn [37–39] which will be conducted at a place of the participants choosing and audio recorded for review. Participants are welcome to have support people with them should they wish. Data will be responses to a range of trigger questions and survey questions to consider the barriers and influences to tobacco, nicotine and cannabis use and cessation, and to understand the population’s health literacy and risk perception related to tobacco, nicotine and cannabis products peri-pregnancy. Participant’s own language and slang will be used when possible. Data will be collected in a coded, but re-identifiable manner to enable researcher and participant follow-up. The code that will be used will be the same as for the participant’s biological and clinical data collection. The codebook will be kept in a separate location to the recording. During the interviews, the participant will only be identified by the unique code.

#### Outcome assessment

Enrollment maternal blood and urine samples provide a baseline and are correlated to exhaled carbon monoxide (CO), cotinine saliva assessment, and the self-reported tobacco, nicotine and cannabis assessment tool (see Figure 4 and Notes: Table 1).

**Table 1.** Research outcomes and statistical analysis.

| **Objective 1:** To accurately describe parental and close household contact(s) peri-gestational patterns of use and exposure to tobacco, nicotine and cannabis products through the creation and use of a validated assessment tool. |  |
| --- | --- |
| Outcomes | Statistical analysis |
| a) Record parental and close house-hold member's patterns of self-reported use and exposure to tobacco, nicotine and cannabis using the NOTICE tool. | a) Descriptive statistics of tobacco, nicotine and cannabis use and exposure self-reported assessment scores will be reported including frequency, means/medians and percentages. The score will be used as a continuous and/or ordinal categorical variable in analyses. |
| b) Conduct biochemical analysis of parental and neonatal samples for tobacco, nicotine and cannabis and metabolites. | b) Descriptive statistics of tobacco, nicotine and cannabis and metabolites levels from collected samples will be reported including frequency, means/medians and percentages. The levels will be used as a continuous and/or ordinal categorical variable in analyses. Participants will be categorised into faster or slower metabolism groups based on their plasma NMR, which is considered the most reliable measure [32, 46]. To ensure the normalization of distribution between the two groups, log-transformed NMR values will be used. In this study, log urine NMR will be utilized across three trimesters of pregnancy, serving as a non-invasive alternative to plasma NMR values. The association between plasma NMR and other NMR values obtained from other biological matrices will be investigated. Two groups (fast and slow metabolisers) will be compared based on their tobacco and nicotine use and exposure behaviours (e.g., cigarettes per day) and TNE levels from maternal urine samples. To assess nicotine exposure in newborn infants, TNE will be calculated for various biological samples including neonate urine, meconium, amniotic fluid, breast milk, and neonate blood samples. A Welch t-test analysis will be used to determine statistically significant differences between the two groups based on their NMR values. Additionally, a regression model will be utilized to present the association between NMR values using different biological matrices. The relationship between plasma NMR values, smoking behaviors, nicotine exposure on newborn babies, and TNE levels will be analysed using a regression model. |
| c) Compare maternal self-reported use and exposure with biochemical assessment of tobacco, nicotine and cannabis concentrations to establish the specificity, sensitivity and predictive validity of the NOTICE tool. | c) The sensitivity, specificity and predictive validity of the screening tool for tobacco, nicotine and cannabis use and exposure will be estimated by comparing self-reported scores with recorded cotinine saliva and CO levels at each antenatal visit.<br>Reliability of the NOTICE tool will also be evaluated using linear regression with urinary nicotine, tobacco and metabolite levels as the outcome and the predictive value of the self-reported scores estimated. |
| **Objective 2:** To determine the pharmacokinetic and pharmacogenomic impacts and outcomes of tobacco, nicotine and cannabis exposure. |  |
| **Outcomes** | **Data and Statistical analysis** |
| a) Investigate specific tobacco, nicotine and cannabis-induced maternal, paternal, foetal, placental and neonatal genomic alternations related to the use and exposure to tobacco, nicotine and cannabis products.<br>b) Determine tobacco, nicotine and cannabis metabolism by genotype (pharmacogenomics).<br>c) Establish the pharmacokinetics of tobacco, nicotine and cannabis in this population. | a, b, c) Descriptive statistics will be used to report the frequency and proportion of participants with identified genetic factors related to the use of tobacco, nicotine and cannabis products. These factors will then be included in univariate and multivariate analyses as being present or absent to estimate their effect on clinical and biochemical outcomes. |
| **Objective 3:** To describe maternal, paternal, placental, foetal and neonatal outcomes according to the use and exposure to tobacco, nicotine and cannabis products and biochemical and genomic analysis |  |
| **Outcomes** | **Data and Statistical analysis** |
| a) Describe maternal, placental, foetal and neonatal outcomes | a) Descriptive statistics of maternal, placental, foetal and neonatal outcomes will be reported including frequency, means/medians and percentages. |
| b) Determine correlations maternal, paternal, placental and neonatal outcomes and parental and foetal | b) Using clinical knowledge and the literature, directed acyclic graphs (DAGs) will be constructed to aid identification of causal effects/associations between 1) clinical outcomes, 2) self-reported tobacco, nicotine and cannabis use, 3) tobacco, |
| tobacco, nicotine and cannabis biochemical concentrations/pharmacokinetics. | nicotine and cannabis metabolite concentrations, and 4) identified biomarkers, genetic/epigenomic/germline alterations, to determine which factors to include in statistical models. |
| c) Determine correlations between maternal, paternal, placental and neonatal outcomes and genomic factors. | c) Multilevel regression modelling will be used to account for the clustering effect (random effects) of each individual mother, to estimate adjusted odds ratios (logistic regression for dichotomous or categorial outcomes) or beta coefficients (linear regression for continuous data outcomes) and 95% CIs for associations. A range of adjustment factors will be considered as both fixed and random effects in the models including gestational age, infant sex, birth weight, preterm delivery, delivery method, post-partum haemorrhage, preeclampsia, maternal age, height, body mass index, parity, urinary tract infection, sexually transmitted infection, diabetes, hypertension, placental data. |
| **Objective 4:** To describe the influences and barriers to cessation for pregnant Australian Indigenous women and their partners or close household contacts to reduce or cease tobacco and nicotine use in pregnancy |  |
| **Outcomes** | **Data and Statistical analysis** |
| a) Qualitative exploration with information rich participants to understand the barriers and influences on tobacco, nicotine and cannabis cessation.<br>b) Understand the population's health literacy and risk perception related to tobacco, nicotine and cannabis products peri-pregnancy. | a & b) Data will be coded and analysis will follow established processes [47] with a preliminary thematic analysis assigning meaning to the data and generating categories and subcategories. The categories most often mentioned will be identified and similarities and dissimilarities detailed. The categories will then be further grouped, and themes and subthemes identified. These will be shared with the research team for review and modification as needed. The consensus themes will be organised for analysis using NVIVO 12. |

At each antenatal visit, maternal urine is collected and correlated to the CO and saliva assessment and the self-reported tobacco, nicotine and cannabis assessment tool. At birth, a second maternal blood is taken to correlate to the excretion of nicotine and cannabis by the mother and neonate as measured in maternal and neonatal urine, breast milk and meconium. The transfer of nicotine and cannabis to the foetus through the placenta and return to the mother is measured by amniotic fluid, arterial and venous cord blood, and placenta samples.

Outcomes include:

1. Parental and close household member’s patterns of self-reported use and exposure to tobacco, nicotine and cannabis using the NOTICE Tool score. Total self-use and total exposure scores will be calculated by summing the total weekly quantity and exposure scores. Scores will then be categorised into clinically meaningful categories, with those reporting no exposure or use categorised as the reference group.
2. Tobacco, nicotine, cannabis and their metabolites concentration in ng/ml or nmol/ml. Concentrations of tobacco, nicotine, trans-3’-hydroxycotinine (3-OH-cotinine), nornicotine, nicotine glucuronide and nicotine N oxide, norcotinine, NNAL, nicotelline, anabasine and anatabine [40, 41] will be summed with the total concentration in each sample recorded as total nicotine equivalents (TNE) and the nicotine metabolite ratio (NMR: 3′-hydroxycotinine/cotinine) will be calculated [42]. Similarly, delta-9-tetrahydrocannabinol-D3 (THC-D3) and delta-8 and/or delta-9 carboxy tetrahydrocannabinol-D3 (THC-COOH-D3) will be measured and summed.
3. DNA methylation patterns influenced by tobacco, nicotine and cannabis use and exposure will be identified. Methylation levels will be assessed either by immunohistological analysis of methylation intermediates (5mC, 5-hmC) or calculated and expressed as β values (β = intensity of the methylated allele (M))/(Intensity of the unmethylated allele (U) + intensity of the methylated allele (M) + 100).
4. Specific tobacco, nicotine and cannabis induced genomic and DNA alterations related to the use and exposure tobacco, nicotine and cannabis products in the different participant groups (maternal, paternal, foetal placental and neonatal). Genotypes influenced by tobacco, nicotine and cannabis use and exposure will be identified. Genome-wide mRNA profiles will be sequenced and expressed as read counts per mRNA.
5. Identified genes and alternations (including gene expression) associated with tobacco, nicotine and cannabis metabolism and pregnancy clinical outcomes. Genes, including CYP2A6 and TCF7L2, with single nucleotide polymorphisms (SNPs) with minor allele frequency greater than 10% will be chosen for inclusion in analyses. PCR results will be reported as negative or positive.
6. Semen volume (mL) and quality including sperm concentrations (million per mL), count (million), progressive motility (%), vitality (%), morphology (%), pH (0-14), leucocyte counts (millions per mL).
7. CO readings will be used as continuous scores and also categorised as: 1) negligible combusted tobacco/cannabis exposure - indicated by <6 CO ppm for non-pregnant adults and <3 CO ppm for pregnant women; 2) light combusted tobacco/cannabis exposure - indicated by 7-10 CO ppm in non-pregnant adults and 4-6 CO ppm for pregnant women; 3) heavy combusted tobacco/cannabis exposure - indicated by >10 CO ppm in non-pregnant adults and >6 CO ppm in pregnant women.
8. Saliva readings will be used as dichotomous scores, categorised as negative or positive
9. Maternal and neonatal outcomes including miscarriage, livebirth/stillbirth, gestational age (weeks), preterm birth (<37 weeks gestation), and birth weight (grams), pregnancy induced hypertension and pre-eclampsia, gestational diabetes, thyroid autoimmune disease, maternal anaemia and other factors of interest are listed in Supplementary Table 1.

### Sample size

Approximately 80 women expecting an Indigenous baby attend antenatal care at Galangoor Duwalami Primary Healthcare Service and the Hervey Bay and Maryborough Hospitals each year, and birth at the Hervey Bay Hospital.

Sample size to validate the NOTICE tool: To estimate the sensitivity and specificity of the NOTICE self-reported assessment tool for tobacco and nicotine use and exposure, assuming tobacco and nicotine exposure and use of 66% [43], a sample of 80 mothers with 5 or more assessments would allow estimation of at least 90% sensitivity and specificity for the screening tool for tobacco and nicotine use and exposure versus urinary cotinine with 95% CI within ±7% of the point estimate [44]. The collection and validation of the additional tobacco and nicotine products increases both the sensitivity and specificity of the tool. Validation will be to POC CO, saliva cotinine, and blood and urine total nicotine equivalents [40, 41]. The collection of cannabis use and exposure measured against urinary cannabinoid concentrations provides an assessment of the tool’s broader utility in pregnancy.

Using multilevel linear regression modelling with urinary cotinine levels as an outcome and adjusted for tobacco and nicotine use and exposure, to estimate a statistically significant change in tobacco and nicotine metabolism during pregnancy in an Australian Aboriginal population, with a power of 80% and alpha level of 0.05, a total of 50 mothers would be required to have a minimum of five tobacco and nicotine urine measurements during pregnancy [45] assuming an intra-class correlation of 0.3 (or lower) between urinary cotinine levels for each mother during pregnancy. With a power of 80% and alpha level of 0.05, 34 placentas would be required to be examined to estimate a 50 cm^2^ difference in area between high and low tobacco and nicotine use/exposure.

### Data analysis

Following data cleaning, checking and validation, the participants’ results from the NOTICE scores for tobacco, nicotine and cannabis use and exposure, the biochemical analysis from all biochemical samples, and the maternal and neonatal outcomes will be examined to address the research objectives as described in Table 1. All data analysis will be conducted using Stata 17 (Statacorp, Texas). Missing data and outliers will be examined and reported. Results will be reported with 95% confidence intervals (95% CI) and statistical significance at alpha 0.05.

## Ethical approval

This project has been approved by the Traditional Owners of the Fraser Coast area, the Butchulla people, in conjunction with Galangoor Duwalami Primary Healthcare Service. The project has the support of the Queensland Aboriginal and Torres Strait Islander Health Council (QAIHC) and has ethics approval from Qhealth (HREC/2021/QRBW/77758) and the University of Queensland (2021/HE002069).

## Discussion

The overarching vision of this clinically derived, clinically driven, community based, mixed method project is to *Close the Gap* in Aboriginal and Torres Strait health outcomes. This project takes a life-course epidemiological approach to health outcomes, focusing on the start of life, that is, maternal and neonatal health to improve whole-of-life health outcomes.

Seventy years of evidence demonstrates that maternal tobacco smoking and exposure to combusted tobacco are the leading modifiable risk behaviours associated with adverse pregnancy outcomes [48]. Currently in Australia, the assessment for tobacco and nicotine exposure during pregnancy is focused on maternal cigarette use - no information on other forms of tobacco, nicotine or cannabis use and exposure is standardly collected or considered to inform clinical care. In addition, fathers or other household members are not asked about their tobacco, nicotine and cannabis use. This limited (or absent) tobacco, nicotine and cannabis screening fails to address the broad range of contemporary products that are used in Australia and thus has ramifications for the mother, the father, the children, the clinician, Indigenous populations, and the broader profile of Australian health.

This project will be reported against the STOBE Guidelines and has purposeful and significant objectives. Firstly, the development of a validated tobacco and nicotine screening tool that can be translated to practice Australia-wide will enhance data reporting and the understanding of tobacco, nicotine and cannabis use and exposure to pregnancy outcomes. In addition, the use of a comprehensive and contemporary screening tool will provide an opportunity for women, families and health professionals to discuss tobacco, nicotine and cannabis use and reduction/cessation options.

Secondly, addressing the absence of literature related to the metabolism of nicotine and the influence of pharmacogenomic factors in Australian Indigenous populations will be transformative. As biotechnology has evolved, there has been an increasing recognition that genetics, epigenetics, and environmental interactions impact on health outcomes. The role of genomics in understanding population risks and targeting prevention or intervention programs to reduce risk or to provide treatment based on genomic knowledge (i.e., a precision medicine approach) is of enormous public health benefit. Genomic profiling allows for the understanding of different outcomes in different populations from the same exposure. Already research exists that shows that nicotine is metabolised differently in genetically different populations [49–53] and particular risks are higher or lower in populations based on these genetic differences, however, this same level of understanding has not been established for Australian Indigenous parental populations. A genome-wide mapping of specific biological samples from the local Australian Indigenous parental population in relation to their potential risk from tobacco and nicotine use and exposure and establishing [the start of] a pharmacogenomic profile will structure a precision medicine approach to health care [54, 55] for this population.

Comprehension and appreciation of the barriers to tobacco, nicotine and cannabis cessation is an essential mechanism in supporting the decrease in tobacco, nicotine and cannabis use. Awareness these factors can lead to the construction of a range of education and support resources which can be selected by future pregnant women and the family to assist them to reduce or cease tobacco, nicotine and cannabis use in pregnancy.

Importantly, the findings from the tobacco, nicotine and cannabis assessment and analysis will be linked to maternal and neonatal outcomes. Using the screening tool as part of standard practice in the future will provide a predictive methodology, enabling expectant mothers, families and health services to better plan birthing and post-birthing needs in situations where tobacco, nicotine and cannabis exposure is an independent factor.

This research project is built on respect for the value of Indigenous perspectives and their contribution to the study. Indigenous knowledge systems are incorporated into the research methodology thereby mutually enriching the research. Translation to practice is an intended outcome of this project but will not be structured until findings are available. The intention is that translation will be informed by the Indigenous participants, the Aboriginal and Torres Strait Islander health service, the research-intensive organisations supporting this research and their researchers, and the chief researcher.

## Limitations

The study consists of some strengths and limitations. One significant strength is the recording of tobacco, nicotine and cannabis use and exposure throughout early to late pregnancy and the collection of a range of biological samples that are used to measure:

- Recency of maternal tobacco and nicotine exposure (maternal CO, saliva, and maternal venous blood and urine),
- The transfer of nicotine to the foetus (venous cord blood, amniotic fluid and neonatal urine),
- The return of nicotine from the foetus (arterial cord blood)
- The longevity of exposure (placenta and meconium)

This approach minimises recall bias and provides a comprehensive and measurable assessment of tobacco and nicotine exposure over the duration of pregnancy. However, the study will only recruit mothers expecting an Australian Indigenous baby in the Fraser Coast area which limits the generalizability of the findings. Additionally, being an observational study, the results will not provide the strongest evidence to establish a causal relationship between nicotine exposure or metabolism and pregnancy outcomes.

## Authors contributions

AR: Conceptualization, design and methodology, establish collaborations and project administration, data collection, resources, writing original draft, review and editing. EAB: Data curation, formal analysis, investigation, methodology, software, validation, writing – review & editing. VB, JB, GM, MS: Conceptualization, supervision, writing – review & editing. GD, SO: Resources, supervision, writing – review & editing. AW: Conceptualization, methodology, resources, supervision, writing – review & editing. SB: Conceptualization, methodology, supervision, writing – review & editing. M-TW: Methodology, writing – review & editing. JM Methodology, supervision, validation, writing – review & editing. KJS: Methodology, resources, supervision, validation, writing – review & editing.

## Data Availability

No datasets were generated or analysed during the current study. All relevant data from this study will be made available upon study completion.

## Acknowledgements

This project could not have developed without the overwhelming endorsement and governance of the Traditional Owners of the Fraser Coast area, the Butchulla people and the Butchulla Aboriginal Corporation and the Butchulla Men’s Business Association. Furthermore, this project cannot progress without the consistent and positive leadership of GD, SO and SB at Galangoor Duwalami Primary Healthcare Service and the engaged involvement of the Galangoor Duwalami teams that wrap around and support the Indigenous expectant families of the Fraser Coast area. Moreover, the cooperation and involvement of maternal services and their support teams from Wide Bay Hospital and Health Services Fraser Coast is essential in ensuring this collaborative project can achieve its aim. Fraser Coast Sullivan and Nicolaides Pathology service are sentinel in the transport of biological samples to Brisbane and the University of Queensland and are providing this service pro bono. In terms of the design, AR conceived, designed the framework of the study, and will lead the data collection. AW, GM, VB, JB and MS guided the data collection design with Indigenous mothers and families and consulted with their respective Indigenous organisations and community members to ensure cultural and community safety and expectations were established. LB designed the statistical analysis and will undertake the data analysis. M-TW will undertake the biochemical analysis as a PhD Scholar at the University of Queensland under the supervision of JM and KS. AR is partially funded under a QHealth Advancing Clinical Research Fellowship.

## Abbreviations

ADHD: attention-deficit/hyperactivity disorder
CO: carbon monoxide
DAGs: directed acyclic graphs
mRNA: messenger ribonucleic acid
nAChR: nicotinic acetylcholine receptors
NMR: nicotine metabolite ratio
NRT: nicotine replacement therapy
POC: point of care
QAIHC: Queensland Aboriginal and Torres Strait Islander Health Council
NOTICE: Ratsch Assessment of Tobacco and Nicotine
SIDS: sudden infant death syndrome
SNPs: single nucleotide polymorphisms
TNE: total nicotine equivalents

## Supporting Information

Supplementary Table 1: Variables of interest for analysis extracted from standard National Perinatal Data Collection report, together with variables of interest for this project (i.e., tobacco, nicotine and cannabis use and exposure)

